# Trends and Patterns of Opioid Epidemic: A Large-Scale Retrospective Study of Hospital Visits with Opioid Poisoning in New York State, 2010-2016

**DOI:** 10.1101/2020.09.01.20185991

**Authors:** Xin Chen, Wei Hou, Sina Rashidian, Yu Wang, Xia Zhao, Richard N Rosenthal, Mary Saltz, Joel H Saltz, Elinor Randi Schoenfeld, George Stuart Leibowitz, Fusheng Wang

**Author notes:** **Corresponding Author:**Fusheng Wang PhD, Department of Biomedical Informatics, Department of Computer Science, Stony Brook University, 2313D Computer Science, Stony Brook, NY 11794-8330.

## Abstract

**Introduction:** To discover trends and patterns of opioid poisoning and the demographic and regional disparities by analyzing large scale patient visits data in New York State (NYS).

**Methods:** Demographic, spatial, temporal and correlation analyses were performed for all OP patients extracted from the New York Statewide Planning and Research Cooperative System (SPARCS) from 2010 to 2016, along with Decennial US Census and American Community Survey zip code level data. The study is based on claims data. 58,481 patients with at least one OP diagnosis and a valid NYS zip code address were included. OP patient counts and rates per 100,000 population; patient level factors (gender, age, race and ethnicity, residential zip code); zip code level social demographic factors. Analyses were completed between 2017 and 2019.

**Results:** In this study of 58,481 patients with opioid poisoning (OP) in New York State from 2010 to 2016, the OP rate increased by 364.6%, and by 741.5% for the age group > 65 years. There were wide disparities among groups by race and ethnicity on rates and age distributions of OP. Heroin and non-Heroin based OP rates show distinct temporal trends as well as major geospatial variation.

**Conclusions:** The findings highlight strong demographic disparity of OP patients, evolving patterns and substantial geospatial variation.

## INTRODUCTION

The United States is experiencing an opioid epidemic and leading in opioid overdose deaths in the world ^1,2^. Opioid overdose can lead to significant morbidity, adverse interpersonal and social impact, chronic diseases, and accidental or intentional death ^3-8^. In 2016, there were more than 42,000 opioid-related deaths in the US ^9^. From 2005 to 2014, opioid poisoning (OP) related hospitalization and emergency room visits increased 64% and 99% respectively ^10^. The number of opioid-related deaths increased more than 4-fold in the United States between 1999 and 2016 ^11^.

Broad demographic measures related to OP rates have been previously reported, but more granularly, the OP problem also has high geographical disparity and impacts different communities to different extents, rendering even county-level data not optimally useful for understanding how to plan appropriate responses to the opioid epidemic. For example, Long Island, which consists of Nassau County and Suffolk County, has been identified as a region with the highest number of opioid overdose deaths in New York State (NYS) ^12^. Such geospatial studies are mainly at state or county level based on OP mortalities, which lacks the spatial resolution to support community level interventions. At the state level, NYS became one of the top 5 states with highest rates of drug overdoses ^13-16^. NYS only publishes county level reports on opioid overdose ^12^.

Given these alarming trends, combating opioid epidemic becomes a high priority for both the US government and State governments such as New York. However, there is a lack of studies to adequately identify, analyze, and monitor the opioid epidemic at the community or patient level. This depends on critical knowledge to answer many questions. For example, which regions or communities have most serious opioid overdose problem and what are the latest trends? What population groups have a higher overdose risk?

As recommended by U.S. department of health and human services, better data and reporting can help us better understand the crisis ^17^. With increased accessibility of health data driven by open data initiatives, large-scale patient level data analysis provides an opportunity for a data-driven approach to identity patterns of the opioid epidemic and to discover potential causes of opioid-related deaths through studying diagnoses from hospital visits at large scale. The NYS Statewide Planning and Research Cooperative System (SPARCS) ^18^ collects patient level details on patient characteristics, diagnoses and treatments, services, and charges for each inpatient stay and outpatient visit for emergency department, ambulatory surgery, and outpatient services. It also includes locations of patients (street addresses). Researchers can benefit from SPARCS data by leveraging patients’ diagnoses, histories and locations. Such unprecedented access to large numbers of patient records linked with spatial data allows us to explore opioid poisoning in NYS with significant improvement of accuracy and coverage compared to prior studies on non-medical opioid use at a national, state or urban level. ^22-23^

Data driven studies from this approach could provide major impact for improving public health studies. Our work on statewide data analytics offers a critical step for community level interventions to prevent or reduce non-medical opioid use and related opioid overdose by identifying high risk communities within which to apply more specific countermeasures. Analyzing opioid data at fine spatial resolution such as zip code can also reveal community level distribution patterns of demographic, regional and temporal trends, which can provide crucial knowledge for local governments, residents, schools, businesses and health systems and service professionals seeking effective solutions to the opioid crisis.

Aiming to better manage the opioid-related public health crisis, we investigated demographic disparities, temporal and geographic patterns of hospital, emergency, ambulatory surgery and outpatient visits (2010-2016) for NYS resulting in at least one OP discharge diagnosis in SPARCS. We performed the following studies to assess: 1) demographics to find disparities of OP rates between age, gender, and race/ethnicity groups; 2) temporal trends of OP rates and any disparities between demographic groups; 3) heroin vs. non-heroin poisoning to understand the differences between two major components of the opioid epidemic; 4) geographic trends of OP rates across NYS at zip code level; and 5) correlations between demographic and socio-economic factors that may be linked to OP.

## METHODS

This study was approved by Institutional Review Board and the Office of Quality and Patient Safety, Department of Health of NYS. Informed consent was not needed as the study had no contact with participants, and the data were obtained from a NYS administrative database. All study analyses were completed between 2017 and 2019.

### Study Populations and Data Sources

In this retrospective cohort study, we used data from SPARCS, an all-payer health claims database for NYS, required for all Article 28 licensed facilities ^18^. We identified OP patients based on ICD codes (primary and secondary diagnosis codes, ICD-9 from January 1, 2010 to September 30, 2015, and ICD-10 from October 1, 2015 to December 31, 2016). Only NYS patients (with a valid NYS 5-digital zip code) with at least one OP related diagnosis were included in analyses. The identified data set included patient level information, such as demographics, diagnoses, treatments, services, addresses of patients, among others.

A list of ICD-9 and ICD-10 diagnosis codes were used to identify the records related to opioid poisoning. The search filter included prior to October 1st, 2015, eight ICD-9 diagnosis codes related to opiates, opium, heroin, methadone, and other narcotics, and after October 1st, 2015, forty-eight relevant diagnosis codes, which were subgroups of topline T400X (opium), T401X (heroin), T402X (other opioids), T403X (methadone) and T404X (synthetic narcotics) ICD-10 diagnostic classes. ^19,20^

### Statistical and spatial analyses

All historical records per individual were aggregated using an encrypted unique patient identifier and ordered by the corresponding hospital visiting dates. For analyzing spatial patterns, the patients were also aggregated by zip code.

For temporal trends, if a patient had multiple OP-related clinical contacts in a year, only the first recorded episode was included. In an analysis combining 7 years’ data, each unique patient was only considered once at their index OP event even if the patient had had multiple OP episodes over more than one year.

OP rates overall and within each sociodemographic category were estimated and compared across 7 years using logistic regression. To evaluate whether OP rates increase linearly over the years, linear trend tests were performed using contrast F tests based on the logistic regression. For correlation studies at zip code level, we used TIGER/Line with Selected Demographic and Economic Data from the US Census Bureau ^24^. To evaluate the correlations between sociodemographic factors with OP rates at zip code level, multiple linear regression models were fit for OP rates as the dependent variable. Zip code level factors, i.e. median household income, race and ethnicity, gender, % of high school education, house units and population density (persons per square mile) were included as covariates. Regression coefficients were standardized in order to compare the effects of factors on different scales.

All maps were generated using ArcGIS Desktop 10.5 (Esri, Redlands CA). Statistical analyses were performed using SAS version 9.4 (SAS Institute, Inc., Cary, NC).

### Patient and public involvement

No patients were involved in setting the research question or the outcome measures, nor were they involved in developing plans for design or implementation of the study. No patients were asked to advise about interpretation or writing up of results.

## RESULTS

Among 146,598,009 encounters of 26,413,181 patients visiting at any facility (inpatient, outpatient, emergency room, ambulatory surgery) in SPARCS, 58,481 patients with an OP-related diagnosis on 72,102 hospital visits between 2010 and 2016 were identified in NYS through the residential zip codes. The overall OP rate of NYS was 296.2 per 100,000 persons over the seven years (Table 1). The counts and rates of OP patients steadily increased during 2010-2016 within each demographic group (all p <0.0001) and for NYS overall (p <0.0001). Each increase overall and within each demographic group demonstrated significant linear trends (all p <0.0001). The comparison between different gender, age, race and ethnicity groups also suggested demographic disparities.

**Table 1.**
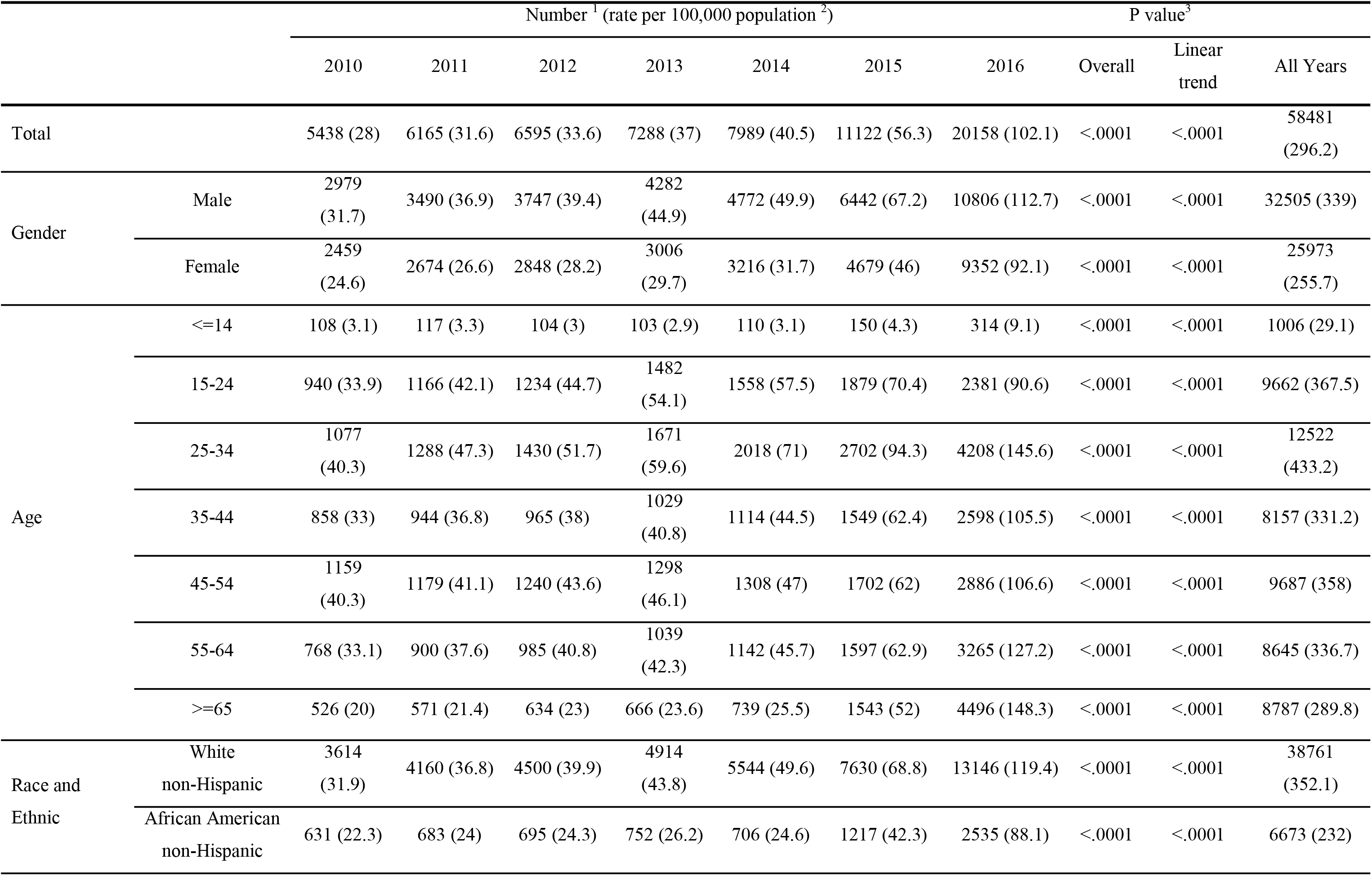
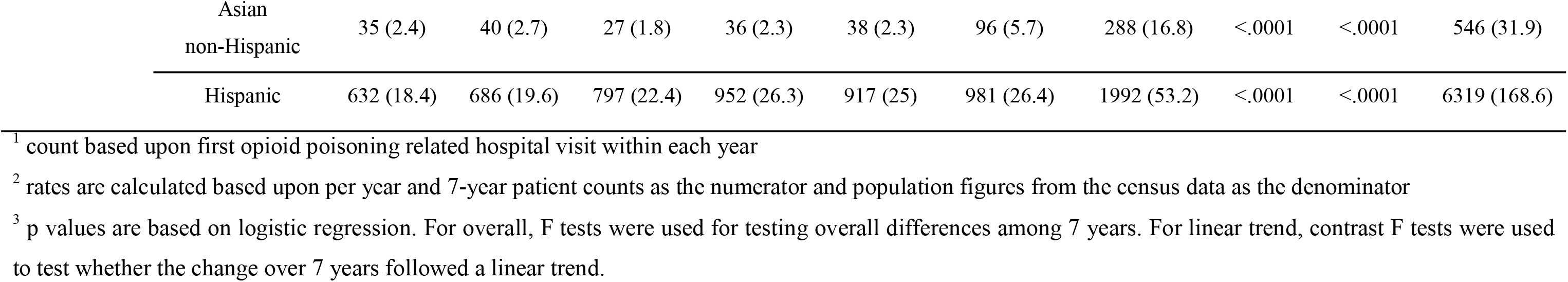
Opioid Poisoning Patient Characteristics in New York State From 2010 Through 2016, Overall and Trends by Years.

### Demographic-based analysis

There were strong disparities among racial/ethnical groups. Figure 1A showed a significantly higher rate of 352.1 per 100,000 among non-Hispanic whites with a significantly lower rate of 31.9 among non-Hispanic Asians (overall p <0.0001). Among different age groups (categorical: 0 to <85 in 5-year increments, and ≥85) for all OP patients in 2010-2016, there was a high peak among young adults (ages in 20-30) and two local peaks among middle ages (ages in 50-60) and babies (ages <5) (Figure 1B).

**Figure 1.**
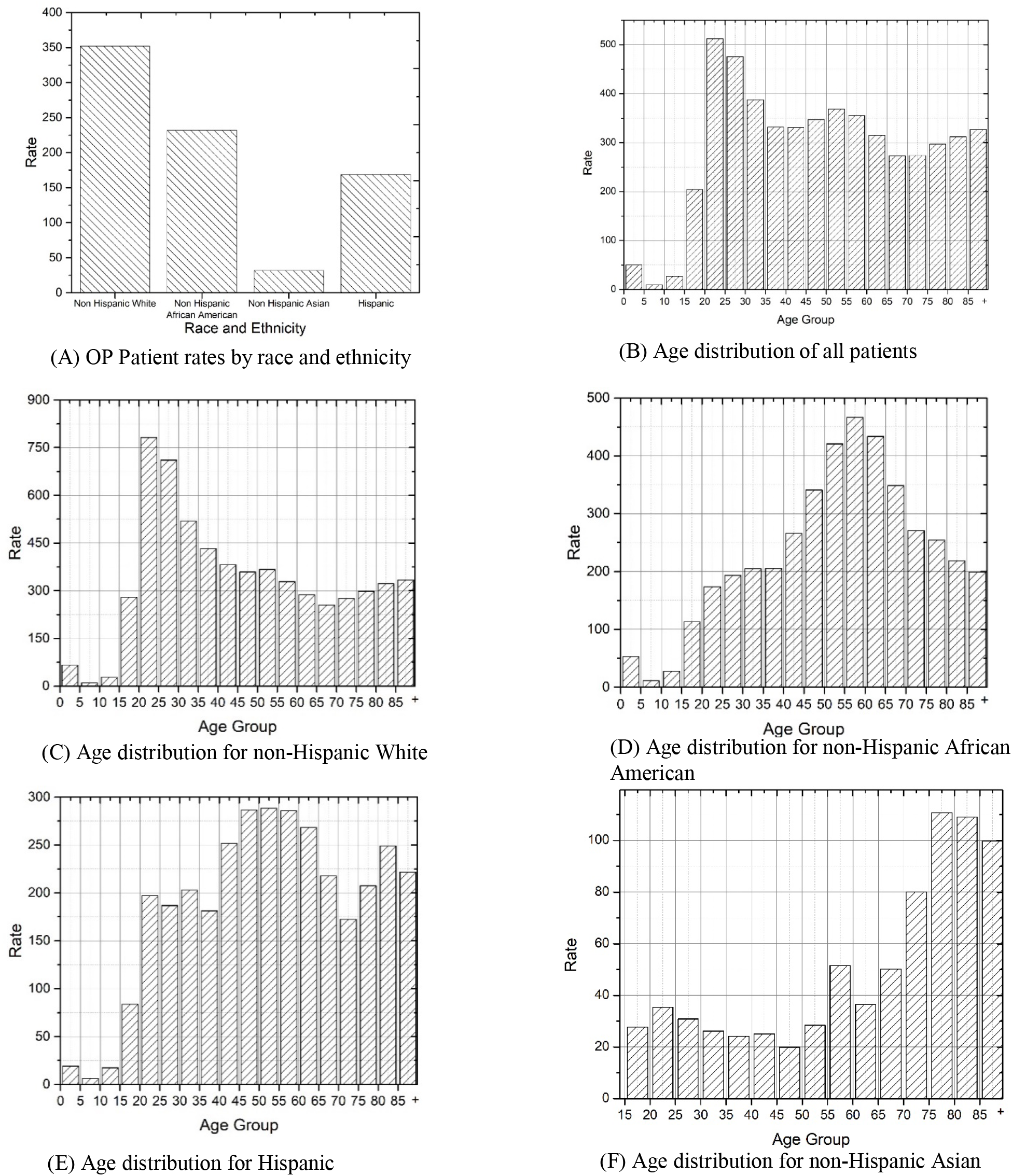
Demographic disparities of opioid poisoning patient rate per 100,000 population, New York State, 2010-2016 The rates are calculated based upon per year or 7-year patient counts as the numerator and population figures from the census data as the denominator.

The age distributions showed significantly different patterns of OP rates between racial/ethnical groups (overall p <0.0001). Non-Hispanic white OP patients were more concentrated among young adults (20-35 age groups) (Figure 1C) whereas the Non-Hispanic African-American and Non-Hispanic Asian OP patients had a higher concentration among 50-70 age groups (Figure 1D) and above 75 age groups (Figure 1F) respectively. There was no obvious peak for the age distributions of Hispanic OP patients with the relatively highest rates among the 45-60 age groups (Figure 1E). The OP rates were not uniformly distributed over age groups within each racial/ethnical group (all p values of goodness of fit test <0.0001, Figures 1C, 1D, 1E and 1F).

### Temporal trends of demographic characteristics

The OP rates for all demographic groups increased dramatically after 2014 (Figure 2). Across years 2010 to 2016, males always have a higher rate than females (all p <0.0001) (Figure 2A). Non-Hispanic whites and non-Hispanic African-Americans increased at a higher rate than Hispanics and non-Hispanic Asians (Figure 2B) (all p <0.0001). The age group of 65+ demonstrated the most dramatic increases after 2014 (Figure 2C) and children (0-14 age groups) demonstrated the slowest rate of increase (p <0.0001).

**Figure 2.**
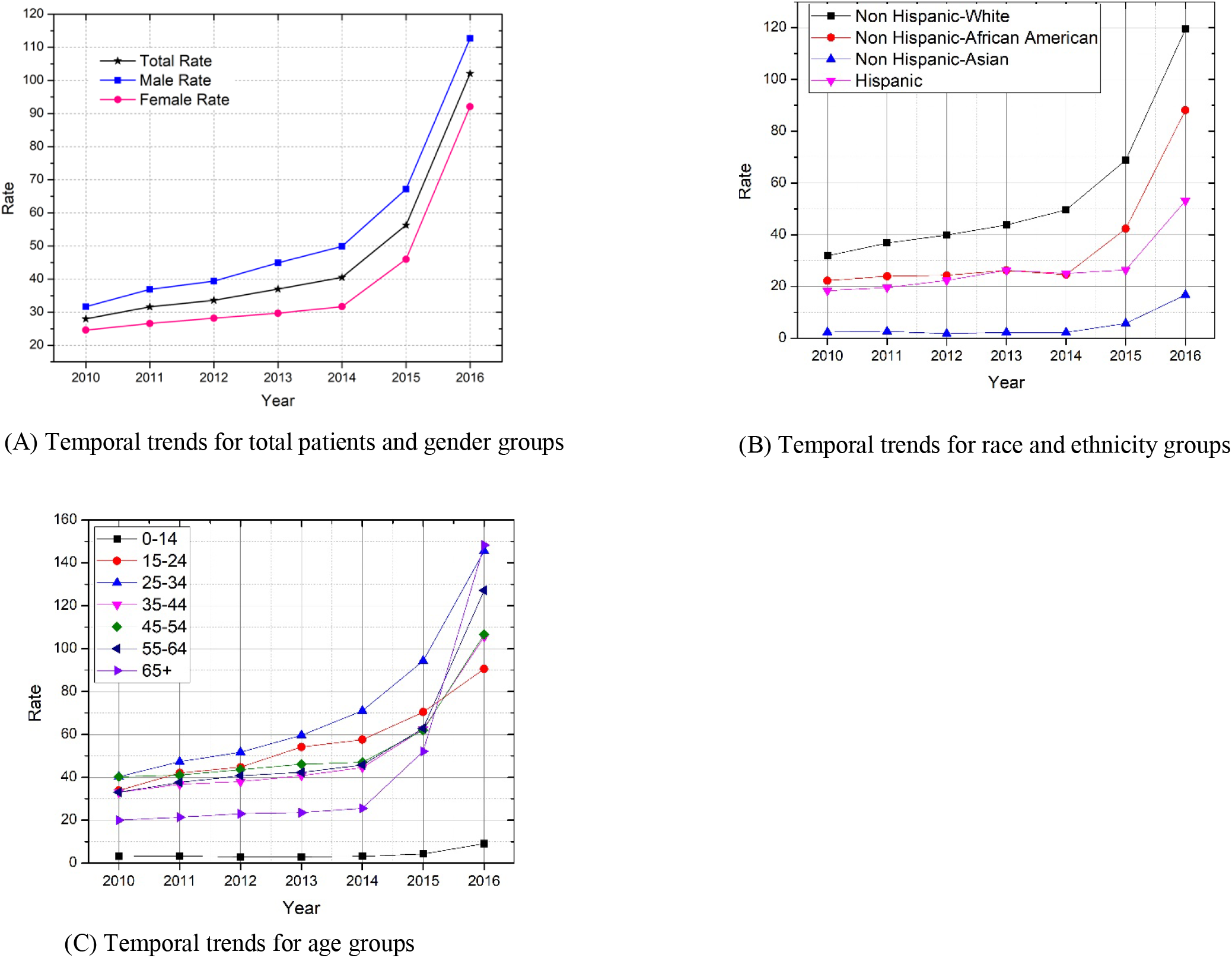
Temporal trends of opioid poisoning patient rate per 100,000 population, New York State, 2010-2016 The rates are calculated based upon per year or 7-year patient counts as the numerator and population figures from the census data as the denominator.

### Comparison between heroin poisoning and non-heroin poisoning

There were strong racial disparities between heroin and non-heroin caused OP patients (Chi-square test p<0.0001, Figure 3A). Non-Hispanic white OP patients had the highest portion attributable to heroin, and most Non-Hispanic Asian OP patients were attributable to non-heroin opioids. The age distribution patterns were distinct for patients with heroin poisoning and those with non-heroin poisoning (Kolmogorov-Smirnov test p<0.0001) (Figure 3B). Young adults within 20-35 age range had the highest OP rates attributable to heroin. Older OP patients were more likely to overdose on non-heroin opioids.

**Figure 3.**
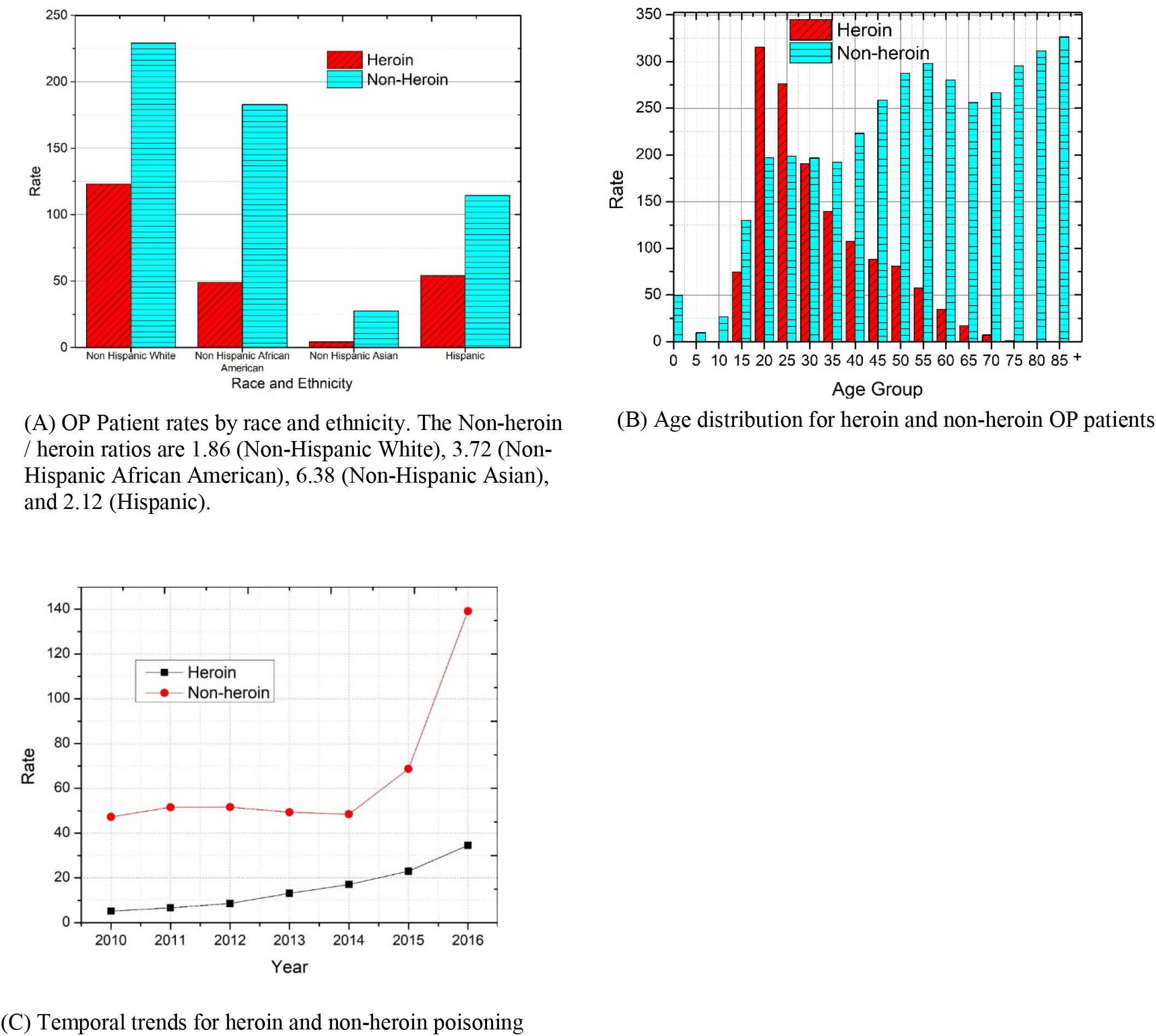
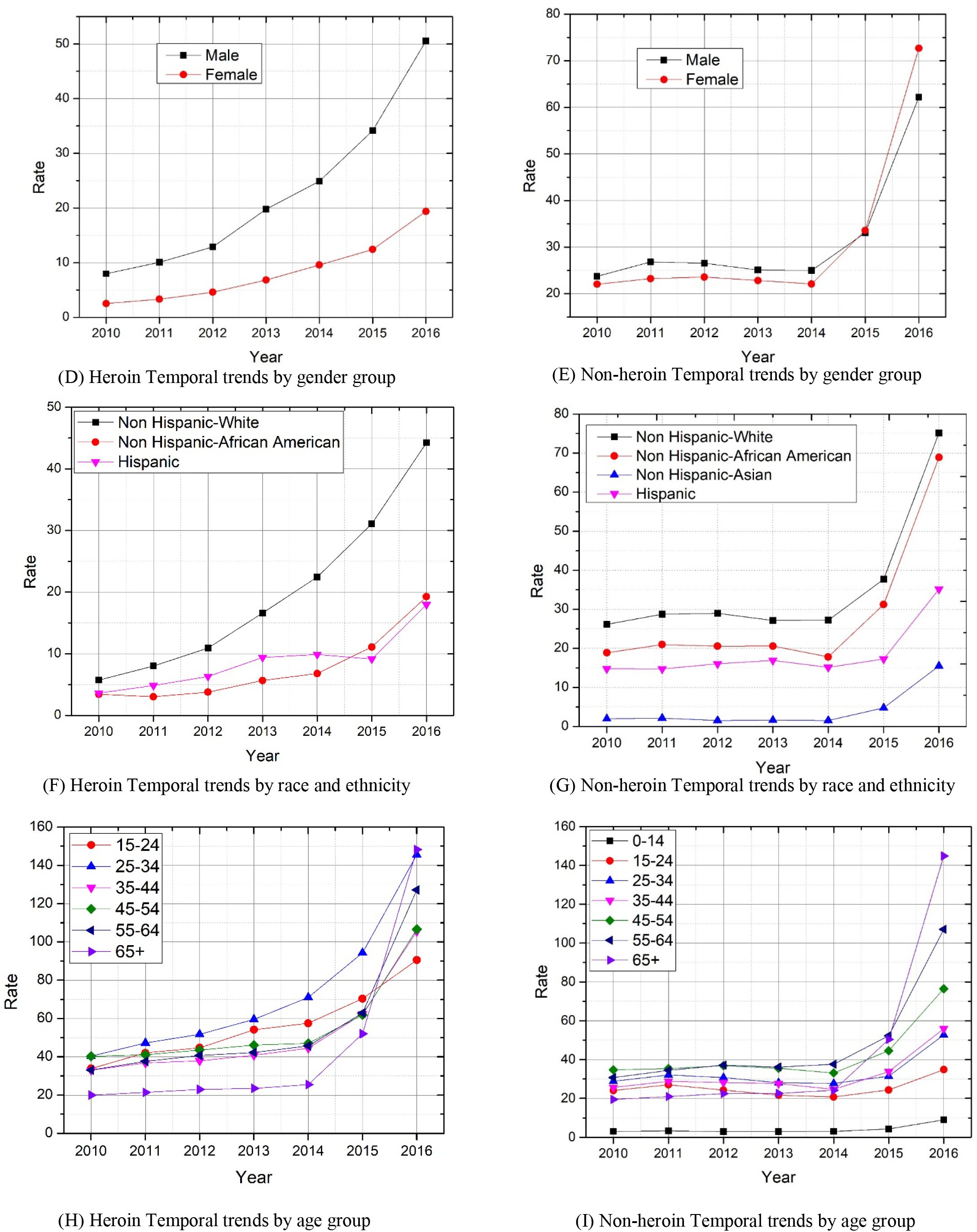
Comparison between heroin and non-heroin related opioid poisoning (OP) patient rate per 100,000 population, New York State, 2010-2016 The rates are calculated based upon per year or 7-year patient counts as the numerator and population figures from the census data as the denominator.

Heroin and Non-heroin based OP groups showed distinct temporal trends (Figure 3C). Year 2015 saw a dramatic increase in the Non-heroin poisoning rate and by 2016, the rate was approximately 4 times that of OP attributable to Heroin (p<0.0001). The rate of heroin poisoning was accelerating in males from 2010-2016 (Figure 3D) and was significantly higher than that of females overall all years (p<0.0001). The rate of Non-heroin poisoning among females began to surpass that of males after 2015 (p for 2015=0.51, p for 2016<0.0001, Figure 3E). The OP rates for Heroin dramatically and consistently accelerated among non-Hispanic whites from 2010-2016 (Figure 3F) and is significantly higher than that of non-Hispanic African American and Hispanic (p<0.0001). The Non-heroin OP rate began to increase after 2014 among all race and ethnicity groups (all p<0.0001) (Figure 3G). For Heroin poisoning, the rate among young adults (25-34) had the most significant increase (Figure 3H). For non-heroin poisoning, the rate of patients among elders (55-64 and 65+) had most significant increase (both p <0.0001) (Figure 3I).

### Geographic patterns of OP counts and rates at zip code level

Appendix Figure 1 presents the maps for both OP rates (Appendix Figure 1A) and counts (Appendix Figure 1B) in NYS in 2016. While major cities (New York, Buffalo, Rochester, Albany) had elevated counts of OP patients (Appendix Figure 1B), New York City (NYC, including the Boroughs of Manhattan, Bronx, Staten Island, and Queens and Brooklyn physically located in westernmost Long Island) had lower OP rates. Long Island (Nassau and Suffolk Counties) had both comparatively high counts and rates. Heroin and Non-heroin based OP mappings demonstrated distinct spatial patterns. Most big cities had higher counts of both heroin and non-heroin OP patients (Appendix Figure 1D, 4F). Higher Non-heroin OP rates (Appendix Figure 1E) were not limited to big cities and appeared across the whole state. An animation video (Video 1) in the Supplemental Content demonstrates the evolving spatial-temporal patterns of OP rates in NYS.

### Sociodemographic Factors Associated with OP

We studied potential correlations between OP rates and demographic and social-economic factors by analyzing a group of demographic and socio-economic factors at the zip code level. Among 8 factors, no high association was found (Supplemental Content, Appendix Table 1). The median household income had a significant but weak negative correlation relationship with overall, heroin and non-heroin OP rates (standardized regression coefficients are −0.235, −0.096 and −0.252 respectively, both p<0.0001), suggesting zip codes with higher OP incidence rate had a lower median household income. Population density (person per square mile) also had a significant but weak negative correlation relationship with heroin OP rates (standardized regression coefficient −0.139, p<0.0001), suggesting zip codes with higher OP incidence rate had a lower population density.

## DISCUSSION

This study provided a large-scale patient level analysis of demographic disparities, spatial patterns, temporal patterns and sociodemographic factors for OP in NYS. With state-wide data including address information, we had sufficient sample size to conduct patient level evaluation at a finer, zip code level of geographic resolution. This study should provide essential new knowledge for local governments and healthcare institutions for design of more focused and targeted interventions.

An important finding was that between 2010 and 2016, the overall rate of OP in NYS increased for patients >65 years by 741.5% compared to the overall 364.6% increase in the general OP rate. This jump is due to the OP rate of 20 per 100,000 in 2010, a relatively low base rate compared to other groups 15-64 years in the same year. Whereas the other racial/ethnic groups had increases of 2.9 to 4 times, Asian, non-Hispanic patients saw a seven-fold increase in the OP rate from 2010 to 2016, with a peak in patients greater than 75. Given that Asian, non-Hispanic patients have a comparatively low prevalence of heroin-associated OP, it appears that patterns of opioid prescribing may be involved in the increased rates of OP in this group from 2010-2016. As such, the study reveals an important finding that can be further explored regarding identifying potential factors such as medical comorbidity (e.g., disorders increasing vulnerability to respiratory depression), differential sensitivity to opioid-induced respiratory depression, and prescribing styles of physicians who treat Asian, non-Hispanic patients. However, we also demonstrate that for the overall group >65 the acceleration in the OP rate is greater than other age groups 15-64 for both Heroin and Non-heroin OP.

### Limitations of this study

There are some limitations inherent to the SPARCS all-payer database from which our analysis derives. Most importantly, there is the potential for systematic over- and under-counting the number of unique individuals who had an episode of opioid poisoning during the look-back period from 2010 to 2016. In 2015, the SPARCS diagnosis codes transitioned from ICD-9 to ICD-10 and might account for a portion of the increased rates observed afterwards ^21^. In particular, the well documented rise in rates of OP related to synthetic opioids such as fentanyl was not captured directly prior to October 2015 as fentanyl and its analogues were not routinely screened for in clinical settings, and the SPARCS identifiers only had a non-specific code of “Poisoning; Other” (Supplemental Content, Appendix Table 2). SPARCS also lacks integration with vital records, thus for example, the records of clinical encounters of OP patients who did not survive prior to arrival at a hospital were not included and may have contributed to a systematic undercount. We intended to further specify the study findings in future work pending incorporating a dataset that has been requested from NYS containing death records linked to SPARCS. For individuals who get treated outside of the SPARCS healthcare provider network, OP events may also have been underreported.

ZIP codes are more likely attributed to roads, post offices, and other facilities within the U.S. postal system, which limited the association between OP incidences and the census related demographic or socioeconomic information.

### Conclusions and relevance

Increasingly availability of open health data provides unique opportunity for a more precise understanding opioid epidemic at large scale. This study demonstrates major disparities among demographic groups and geospatial regions, and evolving patterns of the opioid epidemic in NYS.

## Data Availability

The data is from New York Statewide Planning and Research Cooperative System (SPARCS) which is available for research purposes upon request and approval.

## ACKNOWLEDGMENTS

This work is supported in part by National Science Foundation ACI 1443054 and by NSF IIS 1350885, and Stony Brook University OVPR Seed Grant. The authors disclose no conflicts of interest. The study funders had no role in the design and conduct of the study; collection, management, analysis, and interpretation of the data; preparation, review, or approval of the manuscript; or decision to submit the manuscript for publication.

## APPENDIX

**Appendix Figure 1.**
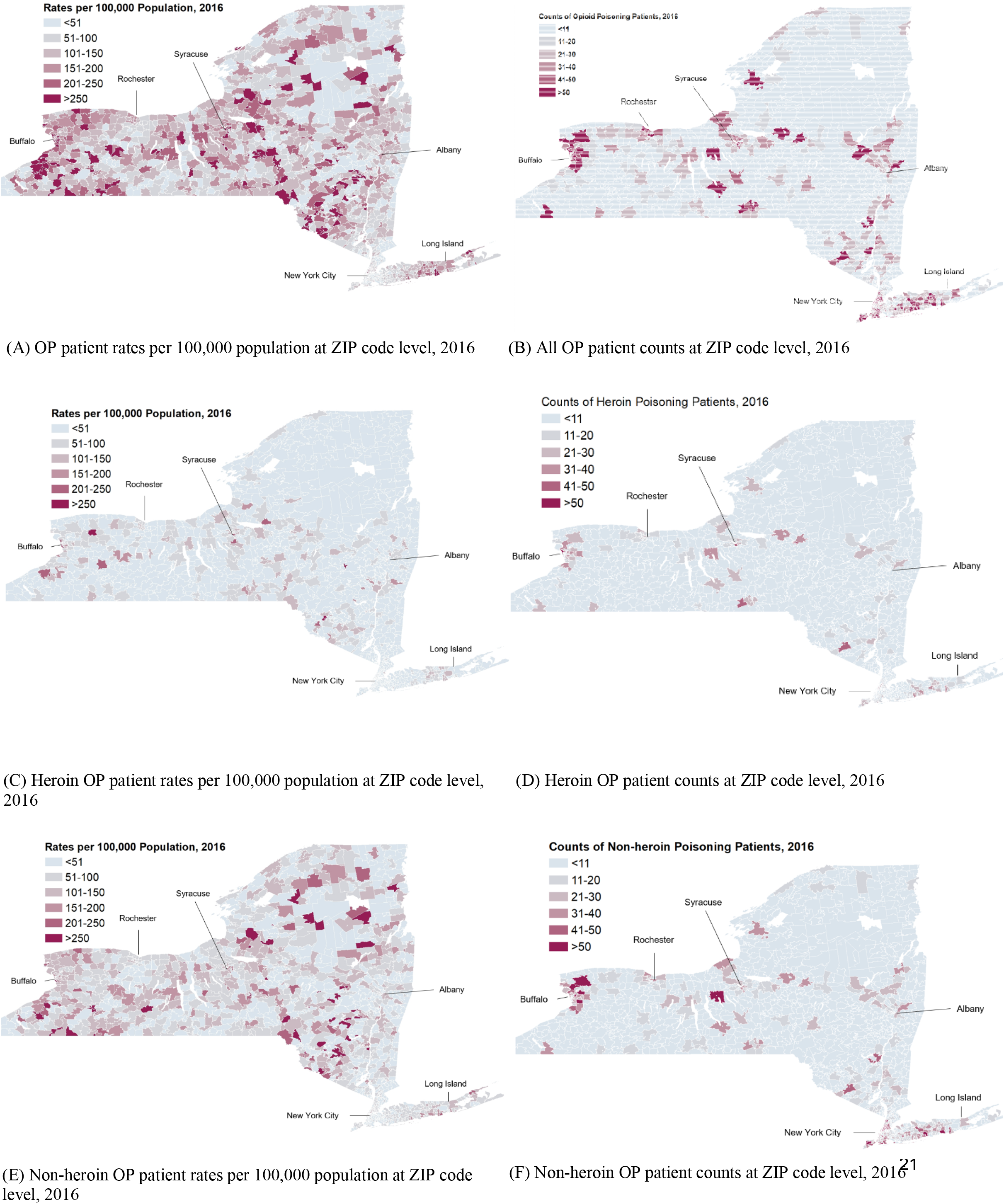
Maps of opioid poisoning (OP) patient counts and rates per 100,000 population at zip code level, 2016.

**Appendix Table 1.**
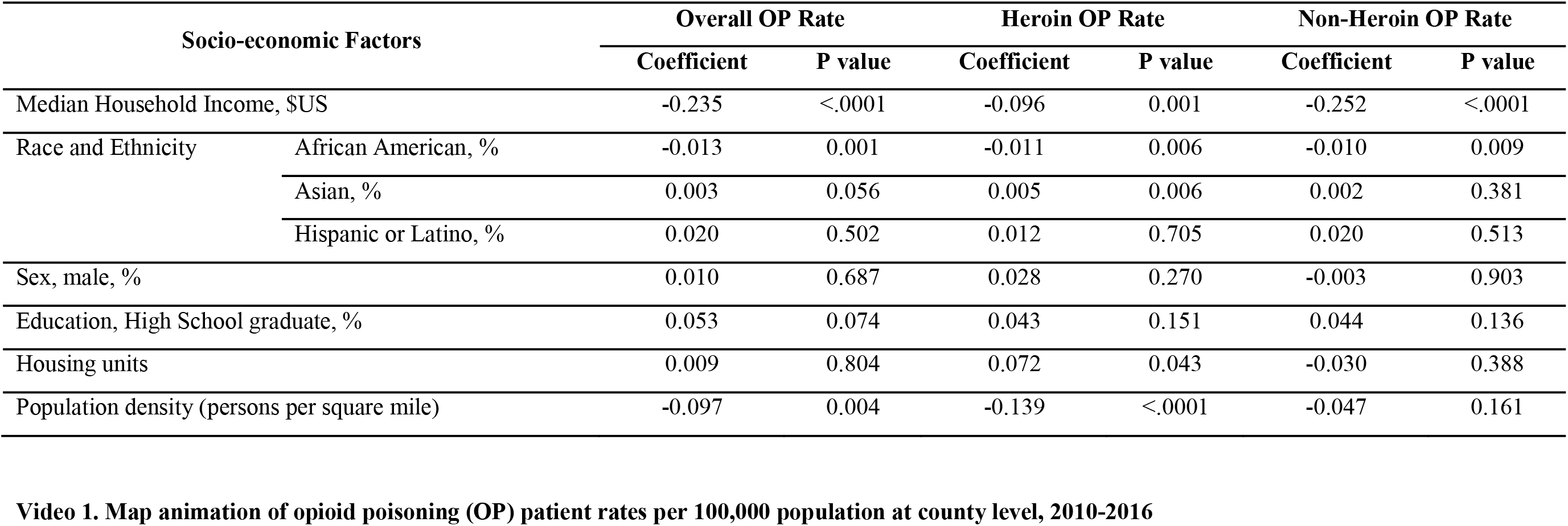
Standardized Regression Coefficients of Social Determinates for Opioid Poisoning Rate per 100,000 population at ZIP Code Level, NYS, 2010-2016.

| Socio-economic Factors |  | Overall OP Rate |  | Heroin OP Rate |  | Non-Heroin OP Rate |  |
| --- | --- | --- | --- | --- | --- | --- | --- |
|  |  | Coefficient | P value | Coefficient | P value | Coefficient | P value |
| Median Household Income, \$US | | -0.235 | <.0001 | -0.096 | 0.001 | -0.252 | <.0001 |
| Race and Ethnicity | African American, % | -0.013 | 0.001 | -0.011 | 0.006 | -0.010 | 0.009 |
|  | Asian, % | 0.003 | 0.056 | 0.005 | 0.006 | 0.002 | 0.381 |
|  | Hispanic or Latino, % | 0.020 | 0.502 | 0.012 | 0.705 | 0.020 | 0.513 |
| Sex, male, % |  | 0.010 | 0.687 | 0.028 | 0.270 | -0.003 | 0.903 |
| Education, High School graduate, % |  | 0.053 | 0.074 | 0.043 | 0.151 | 0.044 | 0.136 |
| Housing units |  | 0.009 | 0.804 | 0.072 | 0.043 | -0.030 | 0.388 |
| Population density (persons per square mile) |  | -0.097 | 0.004 | -0.139 | <.0001 | -0.047 | 0.161 |

**Video 1. Map animation of opioid poisoning (OP) patient rates per 100,000 population at county level, 2010-2016**

**Appendix Table 2.** ICD-9 and ICD-10 Diagnosis Codes Related to Opioid Poisoning. Codes related to heroin are highlighted in bold.

| Years | ICD Code | Version | Description |
| --- | --- | --- | --- |
| 2010-2015 | 9650 | 9 | Poisoning; Opiates and Related Narcotics |
| 2010-2015 | 96500 | 9 | Poisoning; Opium (alkaloids), unspecified |
| **2010-2015** | **96501** | **9** | **Poisoning; Heroin** |
| 2010-2015 | 96502 | 9 | Poisoning; Methadone |
| 2010-2015 | 96509 | 9 | Poisoning; Other |
| **2010-2015** | **E8500** | **9** | **Accidental Poisoning; Heroin** |
| 2010-2015 | E8501 | 9 | Accidental Poisoning; Methadone |
| 2010-2015 | E8502 | 9 | Accidental Poisoning; Other Opiates and Related Narcotics |
| 2015 | T400X1A | 10 | Poisoning by opium, accidental (unintentional initial encounter) |
| 2015 | T400X1S | 10 | Poisoning by opium, accidental (unintentional), sequela |
| 2015 | T400X2A | 10 | Poisoning by opium, intentional self-harm, initial encounter |
| 2015 | T400X2S | 10 | Poisoning by opium, intentional self-harm, sequela |
| 2015 | T400X3A | 10 | Poisoning by opium, assault, initial encounter |
| 2015 | T400X3S | 10 | Poisoning by opium, assault, sequela |
| 2015 | T400X4A | 10 | Poisoning by opium, undetermined, initial encounter |
| 2015 | T400X4S | 10 | Poisoning by opium, undetermined, sequela |
| 2015 | T400X5A | 10 | Adverse effect of opium, initial encounter |
| 2015 | T400X5S | 10 | Adverse effect of opium, sequela |
| **2015** | **T401X1A** | **10** | **Poisoning by heroin, accidental (unintentional), initial encounter** |
| **2015** | **T401X1S** | **10** | **Poisoning by heroin, accidental (unintentional), sequela** |
| **2015** | **T401X2A** | **10** | **Poisoning by heroin, intentional self-harm, initial encounter** |
| **2015** | **T401X2S** | **10** | **Poisoning by heroin, intentional self-harm, sequela** |
| **2015** | **T401X3A** | **10** | **Poisoning by heroin, assault, initial encounter** |
| **2015** | **T401X3S** | **10** | **Poisoning by heroin, assault, sequela** |
| **2015** | **T401X4A** | **10** | **Poisoning by heroin, undetermined, initial encounter** |
| **2015** | **T401X4S** | **10** | **Poisoning by heroin, undetermined, sequela** |
| 2015 | T402X1A | 10 | Poisoning by other opioids, accidental (unintentional), initial encounter |
| 2015 | T402X1S | 10 | Poisoning by other opioids, accidental, sequela |
| 2015 | T402X2A | 10 | Poisoning by other opioids, intentional self-harm, initial encounter |
| 2015 | T402X2S | 10 | Poisoning by other opioids, intentional self-harm, sequela |
| 2015 | T402X3A | 10 | Poisoning by other opioids, assault, initial encounter |
| 2015 | T402X3S | 10 | Poisoning by other opioids, assault, sequela |
| 2015 | T402X4A | 10 | Poisoning by other opioids, undetermined, initial encounter |
| 2015 | T402X4S | 10 | Poisoning by other opioids, undetermined, sequela |
| 2015 | T402X5A | 10 | Adverse effect of other opioids, initial encounter |
| 2015 | T402X5S | 10 | Adverse effect of other opioids, sequela |
| 2015 | T403X1A | 10 | Poisoning by methadone, accidental (unintentional), initial encounter |
| 2015 | T403X1S | 10 | Poisoning by methadone, accidental (unintentional), sequela |
| 2015 | T403X2A | 10 | Poisoning by methadone, intentional self-harm, initial encounter |
| 2015 | T403X2S | 10 | Poisoning by methadone, intentional self-harm, sequela |
| 2015 | T403X3A | 10 | Poisoning by methadone, assault, initial encounter |
| 2015 | T403X3S | 10 | Poisoning by methadone, assault, sequela |
| 2015 | T403X4A | 10 | Poisoning by methadone, undetermined, initial encounter |
| 2015 | T403X4S | 10 | Poisoning by methadone, undetermined, sequela |
| 2015 | T403X5A | 10 | Adverse effect of methadone, initial encounter |
| 2015 | T403X5S | 10 | Adverse effect of methadone, sequela |
| 2015 | T404X1A | 10 | Poisoning by other synthetic narcotics, accidental, initial encounter |
| 2015 | T404X1S | 10 | Poisoning by other synthetic narcotics, accidental, sequela |
| 2015 | T404X2A | 10 | Poisoning by other synthetic narcotics, self-harm, initial encounter |
| 2015 | T404X2S | 10 | Poisoning by other synthetic narcotics, self-harm, sequela |
| 2015 | T404X3A | 10 | Poisoning by other synthetic narcotics, assault, initial encounter |
| 2015 | T404X3S | 10 | Poisoning by other synthetic narcotics, assault, sequela |
| 2015 | T404X4A | 10 | Poisoning by other synthetic narcotics, undetermined, initial encounter |
| 2015 | T404X4S | 10 | Poisoning by other synthetic narcotics, undetermined, sequela |
| 2015 | T404X5A | 10 | Adverse effect of other synthetic narcotics, initial encounter |
| 2015 | T404X5S | 10 | Adverse effect of other synthetic narcotics, sequela |

## Notes

### Competing Interest Statement

The authors have declared no competing interest.

### Author Declarations

The study was approved by the Institutional Review Board of Stony Brook University, and the Office of Quality and Patient Safety, Department of Health of New York State.

## REFERENCES

1. Christie C BC Cooper R Kennedy PJ Madras B Bondi P. The President’s Commision on Combating Drug Addiction and the Opioid Crisis. In. https://www.whitehouse.gov/sites/whitehouse.gov/files/images/Final_Report_Draft_11-1-2017.pdf: White House; 2017.

2. Wonder C. Multiple cause of death 1999-2017.

3. Han B Compton WM Blanco C Crane E Lee J Jones CM. Prescription Opioid Use, Misuse, and Use Disorders in U.S. Adults: 2015 National Survey on Drug Use and Health. Ann Intern Med. 2017;167(5):293–301.

4. American Psychiatric Association. Diagnostic and statistical manual of mental disorders: DSM-5. 5th ed. Washington DC: American Psychiatric Publishing; 2013.

5. Hughes A WM Lipari RN Bose J Copello EAP Kroutil LA. Prescription Drug Use and Misuse in the United States: Results from the 2015 National Survey on Drug Use and Health. In: SAMHSA, ed. Vol September 2016.

6. Schuckit MA. Treatment of Opioid-Use Disorders. N Engl J Med. 2016;375(4):357–368.

7. Durand Z Nechuta S Krishnaswami S Hurwitz EL McPheeters M. Prevalence and Risk Factors Associated With Long-term Opioid Use After Injury Among Previously Opioid-Free Workers. JAMA Network Open. 2019;2(7):e197222–e197222.

8. Oquendo MA Volkow ND. Suicide: A Silent Contributor to Opioid-Overdose Deaths. New England Journal of Medicine. 2018;378(17):1567–1569.

9. Jalal H Buchanich JM Roberts MS Balmert LC Zhang K Burke DS. Changing dynamics of the drug overdose epidemic in the United States from 1979 through 2016. Science. 2018;361(6408):eaau1184.

10. Rudd RA Seth P David F Scholl L. Increases in Drug and Opioid-Involved Overdose Deaths - United States, 2010-2015. MMWR Morb Mortal Wkly Rep. 2016;65(5051):1445–1452.

11. Kiang MV Basu S Chen J Alexander MJ. Assessment of Changes in the Geographical Distribution of Opioid-Related Mortality Across the United States by Opioid Type, 1999-2016. JAMA Network Open. 2019;2(2):e190040–e190040.

12. Health NYSDo. Opioid-related Data in New York State. https://www.health.ny.gov/statistics/opioid/. Accessed 2/15/2018.

13. Centers for Disease Control and Prevention. Drug Overdose Death Data. https://www.cdc.gov/drugoverdose/data/statedeaths.html. Published 2017. Updated 12/19/2017. Accessed 2/15/2018.

14. Hedegaard H WM Miniño AM. Drug overdose deaths in the United States, 1999–2016. In. Vol Data Brief, no 294.. Hyattsville MD: NCHS; 2017.

15. Mack KA JC Ballesteros MF. Illicit Drug Use, Illicit Drug Use Disorders, and Drug Overdose Deaths in Metropolitan and Nonmetropolitan Areas — United States. MMWR Surveill Summ 2017. 2017;66(SS-19):1–12.

16. Volkow ND McLellan TA Cotto JH Karithanom M Weiss SR. Characteristics of opioid prescriptions in 2009. Jama. 2011;305(13):1299–1301.

17. SERVICES USDOHAH. 5-Point Strategy To Combat the Opioid Crisis. 2018.

18. Bureau of Health Informatics. SPARCS Operations Guide. NYS Department of Health; 2016.

19. New York State Department of Health. SPARCS Update. https://www.health.ny.gov/statistics/sparcs/bulletin/jan16.pdf. Published 2016. Accessed 2/12/2018.

20. New York State Department of Health S. Opioid-Related Facility Visits in New York State: Beginning 2010 (Data Dictionary). 2016.

21. Heslin KC Owens PL Karaca Z Barrett ML Moore BJ Elixhauser A. Trends in Opioid-related Inpatient Stays Shifted After the US Transitioned to ICD-10-CM Diagnosis Coding in 2015. Medical Care. 2017;55(11):918–923.

22. Cerda M Gaidus A Keyes KM et al. Prescription opioid poisoning across urban and rural areas: identifying vulnerable groups and geographic areas. Addiction. 2017;112(1):103–112. https://doi.org/10.1111/add.13543.

23. Cerda M Ransome Y Keyes KM et al. Prescription opioid mortality trends in New York City, 1990−2006: examining the emergence of an epidemic. Drug Alcohol Depend. 2013;132(1−2):53–62. https://doi.org/10.1016/j.drugalcdep.2012.12.027.

24. U.S. Census Bureau. TIGER/line shapefiles and TIGER/line files. https://www.census.gov/geographies/mapping-files/time-series/geo/tiger-line-file.html. Published 2018. Accessed December 24, 2019.

25. Schoenfeld, E. R., Leibowitz, G. S., Wang, Y., et al. Geographic, temporal, and sociodemographic differences in opioid poisoning. American journal of preventive medicine, 57(2), 153–164. https://doi.org/10.1016/j.amepre.2019.03.020.

